# Effects of simulated cochlear-implant processing on voice quality distinction: Evidence from analysis of disordered voices

**DOI:** 10.1101/2020.06.29.20142885

**Authors:** Meisam K. Arjmandi, Hamzeh Ghasemzadeh, Laura C. Dilley

## Abstract

The ability to discern variations in voice quality from speech is important for effective talker identification and robust speech processing; yet, little is known about how faithfully acoustic information relevant to variations in talkers’ voice quality is transmitted through a cochlear implant (CI) device. The present study analyzed unprocessed and CI-simulated versions of sustained /a/ vowel sounds from two groups of individuals with normal and disordered voice qualities in order to explore the effects of CI speech processing on acoustic information relevant for the distinction of voice quality. The CI-simulated voices were created by processing the vowel sounds along with 4-, 8-, 12-, 16-, 22-, and 32-channel noise-vocoders. The variations in voice quality for each voice sound was characterized by calculating mel-frequency cepstral coefficients (MFCCs). The effects of simulated CI speech processing on the acoustic distinctiveness between normal and disordered voices were then measured by calculating the Mahalanobis distance (MD) metric, as well as accuracy of support vector machines (SVMs) applied to MFCC features. The results showed that CI speech processing, as simulated by noise vocoding, is highly detrimental to the acoustic information involved in conveying voice quality distinctions. This supports the view that listeners with CIs will likely experience difficulties in perceiving voice quality variations due to the reduced spectral resolution, shedding light on challenges listeners with CIs may face for effective recognition and processing of talkers’ voices.

## 1. INTRODUCTION

The spectro-temporal features in speech contains rich acoustic information from which listeners can learn and retrieve a variety of linguistic and indexical cues that are important for robust speech processing. Voice quality is an important aspect of speech that reflects the configuration and function of individual talkers’ vocal apparatus, contributing important information to speech understanding (Abercrombie, 1967; Podesva, 2007). For example, voice quality variations may provide perceptually salient grammatical and phonological cues for language comprehension (Cameron, 2001; Dicanio, 2009; Dolar, 2006; Garellek & Keating, 2011; Gordon, 2001; Gordon & Ladefoged, 2001; Henton, 1986; Ogden, 2001), not to mention indexical information such as gender, age, and affective state (Abberton & Fourcin, 1978; Laver, 1968; Scott & McGettigan, 2015). Voice quality also contributes to speech understanding through constructing stance in communicative interaction (Aubergé & Cathiard, 2003; Guzman, Correa, Muñoz, & Mayerhoff, 2013; Podesva, 2007; Sicoli, 2010; Tsai et al., 2010; Zimman, 2012).

While listeners with normal hearing (NH) have access to voice quality-related indexical and sociolinguistic information, little is known about how listeners with cochlear implants (CIs) may be disadvantaged in accessing this information due to the reduced spectral resolution. Specifically, it is not clear how the limited spectral resolution in CIs may impact the faithful transmission of acoustic information relevant to the distinction of voice quality. The present work studied voices produced by individuals with normal and disordered voice qualities to examine effects of simulated CI processing on the voice quality distinction.

Acoustic information relevant to voice quality signals a range of attributes, thereby facilitating robust speech perception. This acoustic information allows listeners to associate the variations in talkers’ voice into perceptual attributes of voice quality, such as breathiness, roughness, and strain (Childers & Lee, 1991; Eskenazi, Childers, & Hicks, 1990; Klatt & Klatt, 1990), which can provide acoustic cues for phonemic contrasts (Dicanio, 2009; Garellek & Keating, 2011; Gordon, 2001; Gordon & Ladefoged, 2001). Voicing behaviors like creaky voice signal phrase-final position (Henton, 1986; Ogden, 2001) and convey linguistic information at segmental and prosodic levels (Dilley, Shattuck-Hufnagel, & Ostendorf, 1996; Dilley, Arjmandi, Ireland, Heffner, & Pitt, 2016; Redi & Shattuck-Hufnagel, 2001). These findings support the premise that access to the acoustic information about voice quality facilitates robust perception of speech.

Variations in voice quality also assist listeners in identification of individual talkers’ voices. Multiple traits, such as a talker’s gender, age, dialect, and social group can be often readily incorporated by listeners with NH to recognize talkers’ voice. Voice quality can signal gender (e.g., Gussenhoven, 2004; Ohala, 1983; Puts, Hodges, Cárdenas, & Gaulin, 2007), race (Alim, 2004; Irwin, 1977; Moisik, 2013; Thomas & Reaser, 2004), dialect (Purnell, Idsardi, & Baugh, 1999), and/or social group (Esling, 1978; Sicoli, 2007; Stuart-Smith, 1999)- all of which convey information about talker identity. For instance, studies on African American English demonstrated a connection between talkers’ voice quality and their dialect and race (Arjmandi, Dilley, & Wagner, 2018; Irwin, 1977; Thomas & Reaser, 2004). Non-modal voice qualities were found to be frequently used by African American talkers, leading to a relatively harsh voice quality (Alim, 2004; Britt, 2011). Therefore, perception of acoustic information relevant to voice quality facilities the identification of talkers as an important step in talker normalization toward robust, talker-independent speech perception (Johnson, 2005; Kleinschmidt & Jaeger, 2015; Smith & Patterson, 2005).

Voice quality contributes to learning about several other types of information about talkers such as their physical, psychological and mental health (e.g., voice disorders, anxiety level, mood; e.g., Eskenazi et al., 1990; Kreiman, Vanlancker-Sidtis, & Gerratt, 2005). Vocal fold disorders (e.g., vocal fold polyps, nodules, etc.) represent an instance where voice quality is abnormally altered (Arjmandi & Pooyan, 2012; Arjmandi, Pooyan, Mikaili, Vali, & Moqarehzadeh, 2011; Ghasemzadeh & Arjmandi, 2019; Umapathy, Krishnan, Parsa, & Jamieson, 2005). These abnormalities (physiological, neurological, and/or functional) are associated with the variations in talkers’ voice quality and are recognized as disordered voice quality, as compared to normal voice quality (Blood, Mahan, & Hyman, 1979; Oates, 2009). Abnormalities associated with voice disorders lead to perturbations of vocal fold vibratory patterns in some or all glottal cycles, which can impact voice spectra in multiple frequency regions (Eskenazi et al., 1990; Hammarberg, Fritzell, Gaufin, Sundberg, & Wedin, 1980; Hartl, Hans, Vaissière, Riquet, & Brasnu, 2001; Krom, 1995; Naranjo, Lara, Rodríguez, & García, 1994; Wolfe, Cornell, & Palmer, 1991). Examples of acoustic manifestations of disordered voices include changes in the low and high frequency energy profiles of talkers’ voices (e.g., increased spectral energy within the mid- and high-frequency regions) (Arjmandi & Pooyan, 2012; Ball & Code, 2008; Ghasemzadeh & Arjmandi, 2019; Kitzing & Åkerlund, 1993; Naranjo, Lara, Rodríguez, & García, 1994), as well as decreased steepness of the low-frequency spectral slope (Arjmandi & Pooyan, 2012; Behroozmand & Almasganj, 2007; Hartl et al., 2001; Klatt & Klatt, 1990; Guus de Krom, 1995). Taken together, these studies show that disordered voice quality impacts spectral characteristics of voice in multiple sub-bands in ways that are distinctive from spectral profile of normal voice.

Unlike listeners with NH who have access to acoustic information signaling voice quality, access to spectral profiles of voice through CIs is limited by factors such as the number of spectral channels (cf. number of electrodes) and mismatches between the frequencies in acoustic content and frequencies associated with auditory nerves at the place of electrode stimulation on the cochlea (e.g., Fu, Chinchilla, Nogaki, & Galvin, 2005; Svirsky, 2017). As a result, listeners with CIs receive partial, degraded representations of acoustic information in talkers’ voices. An important portion of voice quality-related acoustic information is associated with the change in vocal folds that appears in mid- and high-frequency regions of the voice spectrum (Arjmandi & Pooyan, 2012; de Krom, 1993; Fukazawa, el-Assuooty, & Honjo, 1988; Hartl et al., 2001; O’Leidhin & Murphy, 2005; Yumoto, Gould, & Baer, 1982). This acoustic information includes, but is not limited to, elevated energy levels in these frequency regions and deformation of formant frequencies. These spectral regions are either completely filtered or partially transmitted through CIs (Goupell, 2015; Svirsky, 2017). Low-frequency harmonics are also affected through CIs. Acoustic cues such as fundamental frequency (*F_0_*) and *H1-H2* (first and second harmonic amplitude differences) and *H1-A1* (first harmonic and first formant amplitude differences) are low-frequency harmonic components that moderately reflect changes in the quality of talkers’ voice (e.g., breathy, strained, rough, etc.), where these can potentially contribute to recognition of talkers’ gender and individual identity (Fu, Chinchilla, & Galvin, 2004; Gelfer & Bennett, 2013). We currently have limited evidence about how the reduced spectral resolution in CIs may degrade acoustic information relevant to detecting voice quality variation.

In the present study, we used voice samples from two groups of talkers, one with normal voice quality and one with disordered voice quality, in order to examine effects of simulated CI processing of such voices on acoustic information available for distinguishing between these two classes of voice qualities. Noise-vocoding was used to simulate the limited spectral resolution of CIs in representation of the acoustic information through varying the number of spectral channels in the noise vocoder, thereby simulating varying degrees of spectral resolution while keeping other parameters of the simulation constant (e.g., the slopes of the synthesis bands). Acoustic simulation of CI speech is a useful and precious approach to simulate what acoustic information listeners with CIs have access to when hearing speech (Do, 2012; Do, Pastor, & Goalic, 2012; Santos, Cosentino, Hazrati, Loizou, & Falk, 2013). In our analysis, we first investigated the variations in the magnitude spectrum of normal and disordered voices to understand how CIs may impact reception of information pertinent to distinguishing between these two classes of voice qualities. We further quantified the acoustic distance between normal and disordered voices as a function of the number of spectral channels, in order to examine effect of CI-related spectral resolution (number of channels in the noise vocoder) on availability of acoustic information related to voice quality distinctions.

## 2. MATERIALS AND METHODS

### 2.1. Voice samples

The voice samples in this study were sustained /a/ vowel sounds from the voice disorders database model 4337, version 1.03 (Kay Elemetrics Corporation, Lincoln Park, NJ), developed by Massachusetts Eye and Ear Infirmary (MEEI), Voice and Speech Lab. Two groups of participants who were diagnosed as having either normal or disordered voices were asked to sustain the vowel /a/. Participants’ voices were recorded at a sampling frequency of 44.1 kHz with 16-bit resolution. All the analyzed voice segments were 1-second long, extracted from the middle of each excerpt to deal with the length difference between normal and disordered voice samples, as well as the transient patterns during the onset and offset of the phonations. The vowel sounds from 293 individuals were analyzed. 53 talkers had normal voice quality (21 males) and the remaining 240 talkers (96 males) were diagnosed with one or multiple voice disorders (e.g., vocal fold nodules, vocal fold paralysis, vocal fold cysts) that had resulted from abnormal physiological, neurological, and/or functional changes affecting vocal fold health.

### 2.2. Creation of noise-vocoded voice samples

CI-simulated versions of unprocessed voice samples were created using a noise-excited envelope vocoder, the AngelSim^TM^ Cochlear Implant and Hearing Loss Simulator (Fu, 2019; Emily Shannon Fu Foundation, www.tigerspeech.com). The CI-simulated vocoding simulation process used in prior studies was followed to create CI-simulated voices (Shannon, Zeng, Kamath, Wygonski, & Ekelid, 1995). This process involved dividing each voice spectrum into a variable number of logarithmically-spaced frequency bands between absolute lower and higher frequencies of 200 Hz and 7000 Hz (24 dB/octave analysis filter slopes), corresponding to the frequency-place map simulated by the Greenwood function (Greenwood, 1990). These frequency limits approximate the corner frequencies in Cochlear Nucleus speech processors (Crew & Galvin, 2012; Winn & Litovsky, 2015). The amplitude envelope of each signal, obtained from filtering the voice spectrum under each sub-band, was captured using half-wave rectification and a low-pass filter with a cut-off frequency of 160 Hz and filter slope of 24 dB/oct; this simulated the performance of the average CI listener in envelope discrimination (Chatterjee & Oberzut, 2011; Chatterjee & Peng, 2008). The extracted amplitude envelopes were then used to modulate band-pass filtered white-noise carrier signals, which were created using a filter identical to that implemented for the analysis filter. The final noise-vocoded version of each voice stimulus was created by summing amplitude-modulated signals. This process replaces fine spectro-temporal structures in voice signal with noise while preserving most of the coarse-grained temporal structures. The quality of CI-simulated voice depends on the number of spectral channels in the vocoder. The noise-excited envelope vocoder was used in *AngelSim* software to process unprocessed voices and create their noise-vocoded versions with 4-, 8-, 12-, 16-, 22-, and 32-channel. Therefore, the simulated cochlear-implant voices were created at six levels of spectral degradation (4-, 8-, 12-, 16-, 22-, and 32-channels). The choice of the number of spectral channels was made to simulate a wide range of spectral degradation and their corresponding perceived difficulty in speech processed through CIs (Shannon, Fu, & Galvin, 2004), as well as to cover the current set-up of between 12-24 active channels in cochlear implant devices. Considering the assumption made about the relationship between electrical spread in the cochlea and acoustical filter slope (Bingabr, Espinoza-Varas, & Loizou, 2008; Oxenham & Kreft, 2014), the selected filter slope of 24 dB/Oct is in the highest range of steepness provided by current CI technology (filter slope varies between 8 and 24 dB/oct), corresponding to the minimum channel interaction available in the current CI devices.

### 2.3. Analysis of voice spectra

We first investigated the average spectra of the two groups of voices (unprocessed normal voices vs. unprocessed disordered voices) to understand how the noise-vocoder and the number of spectral channels would likely affect the distinctive features of these two classes of voice qualities across frequency regions. The average spectrum of voice signals derived over the spectrum of all voice samples from each class of voice quality (i.e., normal or disordered) were estimated using 12^th^-order linear predictive coding (LPC) (Rabiner & Schafer, 1978). Variations in characteristics of average voice spectra were investigated under seven levels of spectral degradation (unprocessed, 32, 22, 16, 12, 8, and 4-channels noise-vocoder) to simulate how differing fidelity of CI processing may affect the acoustic information available to signal the distinction between normal vs. disordered voice qualities.

### 2.4. Using MFCC features to characterize acoustic information

Mel-frequency cepstral coefficients (MFCCs) were used to characterize variations in spectral profile of unprocessed normal vs. disordered voices, as well as their CI-simulated counterparts. MFCC features approximate the filtering structure and frequency resolution of the human auditory system (Fant, 1973; Hunt, Lennig, & Mermeletein, 1980; Davis and Mermelstein, 1980; Shaneh & Taheri, 2009; Stevens, Volkmann, & Newman, 1937). MFCC features have been shown in prior studies to characterize well canonical features distinguishing normal and disordered voices (Ali, Alsulaiman, Muhammad, Elamvazuthi, & Mesallam, 2013; Dibazar, Narayanad, & Berger, 2002; Firdos & Umarani, 2016; Godino-Llorente, Gomez-Vilda, & Blanco-Velasco, 2006; Panek, Skalski, Gajda, & Tadeusiewicz, 2015). Furthermore, features that are developed based on speech production behaviors such as *F_0_* and the first and second harmonics (*H1* and *H2*) are either heavily degraded or are absent in the CI-simulated voices. Using MFCC features also permitted us to deal robustly with these methodological challenges on the way to characterizing changes in spectral properties of talkers’ voice spectra across normal vs. disordered voice quality.

Fig. 1A shows the schematic of the approach used for calculating MFCCs for samples of voices with normal and disordered qualities. To calculate MFCCs, each /a/ vowel sound (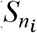 or 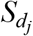 in Fig. 1A) was first segmented into frames of 30 ms with a frame shift of 15 ms. Here, *i* and *j* indicate the index of voice stimuli for normal and disordered voices, respectively (*i* = {1,2,3, …,53} and *j* = {1,2,3, …,240}). A Hamming window was then applied to each frame to decrease the effect of sidelobes for better frequency-selective analysis (Rabiner & Schafer, 1978). The power spectrum of each frame was calculated based on Fast Fourier transform (FFT) analysis. Then, 32 mel-filterbanks were generated and applied to voice power spectra. MFCCs were derived by calculating the DCT of the logarithm of all filterbank energies (Rabiner & Schafer, 1978). Eventually, the first twelve components were preserved as MFCC features for each frame of a voice signal (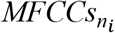 or 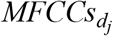 in Fig. 1). Each voice signal was represented by an MFCC matrix of size *F*×*12*, where *F* indicates the number of frames in a voice signal. The same procedure was performed to calculate MFCC matrices for CI-simulated versions of the unprocessed normal and disordered voices, as indicated by dashed lines in Fig. 1A.

**Fig. 1.**
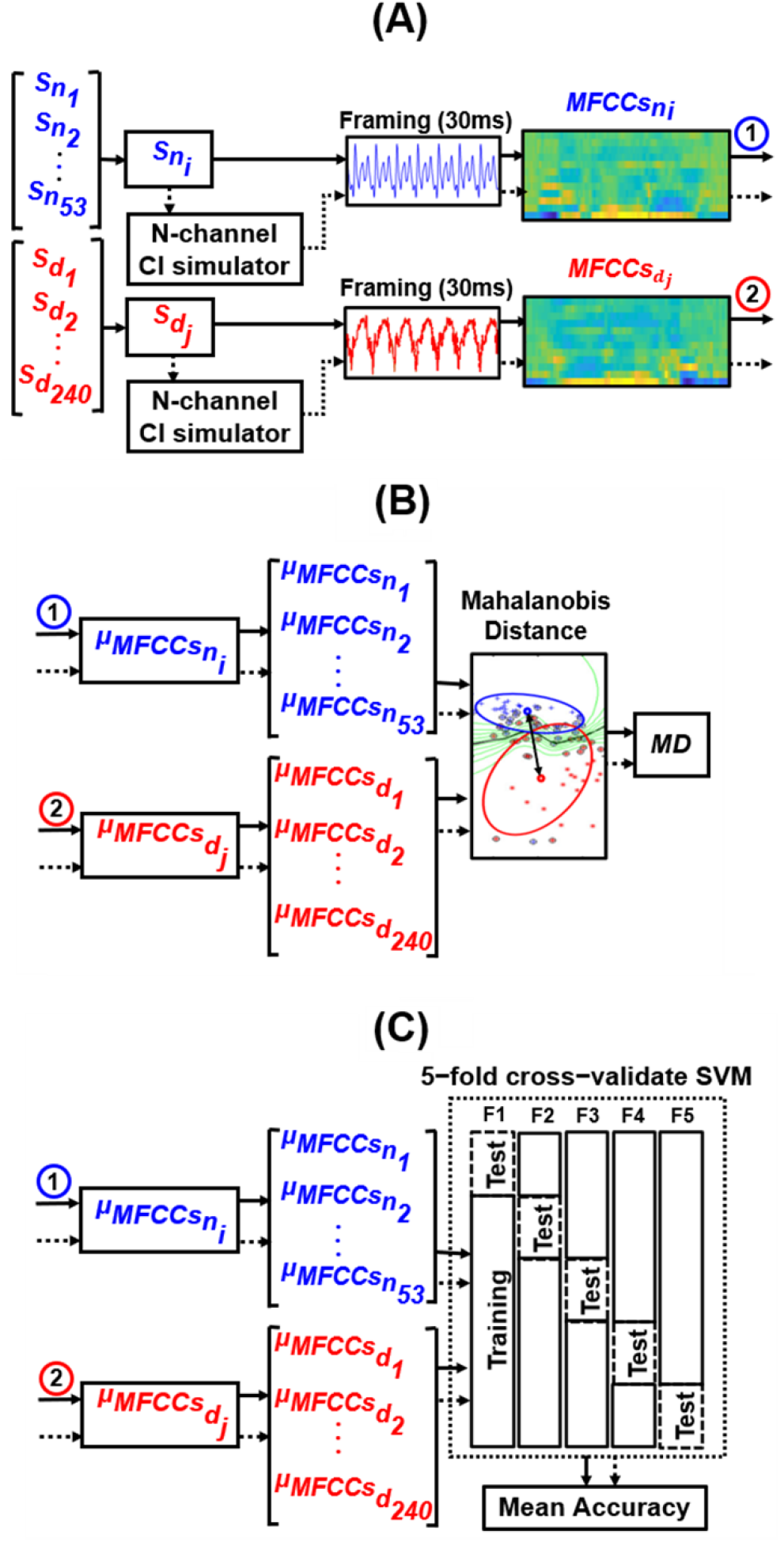
(Color online) Schematic diagram of methods used in the current study to (A) characterize acoustic properties of normal and disordered voice stimuli based on MFCCs features; (B) evaluate the acoustic distance between voices with normal and disordered qualities based on calculating *Mahalanobis distance* between MFCCs; and (C) evaluate the acoustic distance between normal vs. disordered voice qualities based on the classification accuracy derived from applying 5-fold validation to a support vector machine (SVM) classifier. The dashed lines refer to the process for creating and analyzing the CI-simulated versions of the voice stimuli. N in the “*N-channel Simulator*” block stands for the number of spectral channels in the CI-simulated noise vocoder. Components, blocks, and lines with blue (dark gray) color show the paths for processing voices with normal quality, whereas the components with red (light gray) color show the paths for processing voices with disordered quality.

### 2.5. Acoustic distance quantification using Mahalanobis distance

To examine the acoustic distance between two classes of normal and disordered voices as a function of the level of spectral degradation (imposed by the variable number of spectral channels in the CI-simulated voices), we calculated Mahalanobis distance (MD) metrics on MFCC features. MD is a distance measure, which calculates the distance between two or more classes at a multidimensional feature space (Arjmandi et al., 2018; Maesschalck & Massart, 2000; Masnan et al., 2015; Xiang, Nie, & Zhang, 2008). MD is analogous to a multidimensional d’, as used in signal detection theory (Macmillan & Creelman, 2004). This multivariate statistical approach uses two feature matrices (or vectors) from two separate classes to evaluate the extent to which the two classes can be distinguished after sphering the distance matrix between the two classes using the average covariance matrix of the per-class centered data (Maesschalck & Massart, 2000; Masnan et al., 2015). Hence, a relatively greater MD value for a condition (e.g., unprocessed) means a relatively larger distance between the two classes of normal and disordered voice qualities with a relatively lower between-class overlap for that condition compared to other conditions (e.g., 32-channel noise-vocoded voices). Therefore, a relatively larger distance indicates the presence of more discriminative acoustic information relevant to the distinction between two classes of voice qualities for a condition compared to other conditions.

As shown in Fig. 1B, the acoustic properties of each voice stimulus (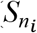 or 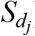) were characterized by a single, time-averaged MFCC vector (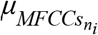 or 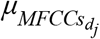), derived by averaging MFCCs across frames of each voice stimulus. These time-averaged MFCC features have been shown to successfully represent the unique spectral characteristics of a speech sound (Mckinney & Breebaart, 2003; Davis and Mermelstein, 1980; Terasawa, Slaney, & Berger, 2005). Therefore, 53 normal voices were presented by fifty-three 12-dimensional average MFCC vectors, constructing a 53×12 feature matrix. Likewise, 240 disordered voices were characterized by 240 12-dimensional average MFCC vectors, leading to a 240×12 feature matrix. The acoustic distance between normal and disordered voices was then measured by calculating the MD between these two matrices of MFCCs (Fig. 1B). The same procedure was followed for simulated versions of the unprocessed normal and the unprocessed disordered voice stimuli (dashed lines in Fig. 1B), leading to seven values of MD corresponding to seven levels of spectral degradation from the unprocessed to 4-channel noise-vocoded voices. The calculated MDs of seven levels of spectral degradation were comparatively examined to identify the extent to which the spectral information involved in distinguishing voices with normal and disordered qualities were affected by CI noise vocoding, as well as the number of channels in the CI noise-vocoding process.

### 2.6. Acoustic distance quantification using support vector machines (SVMs)

We further used SVMs classifier to measure the acoustic distance between two classes of voice qualities. SVMs have been frequently used as a successful classification method for various classification purposes, including classification of normal and disordered voice qualities (Akbari & Arjmandi, 2015; Arjmandi et al., 2011; Ghasemzadeh, Khass, Arjmandi, & Pooyan, 2015; Ghasemzadeh & Arjmandi, 2019; Arjmandi, Pooyan, Mohammadnejad, & Vali, 2010; Umapathy, Rachel, & Thulasi, 2018). One application of SVM classifiers is to evaluate features that maximally distinguish between two classes, where the classification accuracy of SVM classifiers is a criterion for feature evaluation (Heijden, Ferdinand, Ridder, & Tax, 2005). We took advantage of this property of SVMs to examine effects of CI speech processing and number of channels on the acoustic distinctiveness between normal and disordered voices, providing a complementary analysis to MD. Higher classification accuracy between two classes indicates that there was more distinctive acoustic information with respect to class separation.

As illustrated in Figure 1C, a 5-fold cross-validation analysis was performed to train and then test an SVM classifier on its classification accuracy in distinguishing between normal and disordered voice qualities at seven levels of spectral degradation (unprocessed, 32-, 22-, 16-, 12-, 8-, and 4-channel). Two feature matrices of 53×12 and 240×12 MFCC features from normal and disordered classes were entered into the SVM classifier for training and testing phases, as executed through the 5-fold cross-validation procedure (Kohavi, 1995; Reilly, Moran, & Lacy, 2004). The output of the SVM classifier was the mean SVM classification accuracy over classification accuracies, obtained from five repetitions of cross-validation. The average classification accuracies at six levels of spectral degradation were examined with reference to that of the unprocessed condition (as baseline performance) to understand the extent of degradation imposed by CI noise vocoding on acoustic information involved in voice quality distinction. The radial basis function (RBF) kernel was used in SVM classifier. The parameters of RBF kernel and the regularization parameter (ξ) of SVM were set to their default values in *Matlab*.

## 3. RESULTS

In three steps, we examined the effects of simulated cochlear-implant processing on spectral information relevant to voice quality distinction. We first examined spectra of normal and disordered voice samples under the seven spectral degradation conditions. Fig. 2 shows the average magnitude spectra of the two groups of voices with normal (blue or dark gray) and disordered (red or light gray) qualities across all voice samples. The standard deviations of magnitude spectra are also shown as a shaded area across the average lines. The average voice spectra are selectively shown for unprocessed (panel A) and simulated cochlear implant voices with 16- (panel B) and 4-channel (panel C) spectral resolution. These spectra are computed by averaging individual frequency spectrum over all voice samples from a class of voice quality. Overall, these plots demonstrate the detrimental effect of CI processing on spectral information that could signal differences in talkers’ voice quality. The patterns of variation in average spectral energy of disordered voices compared to normal voices at different frequency sub-bands reflect voice quality variations, caused by a wide range of physiological, neurological, and/or functional voice disorders.

**Fig. 2.**
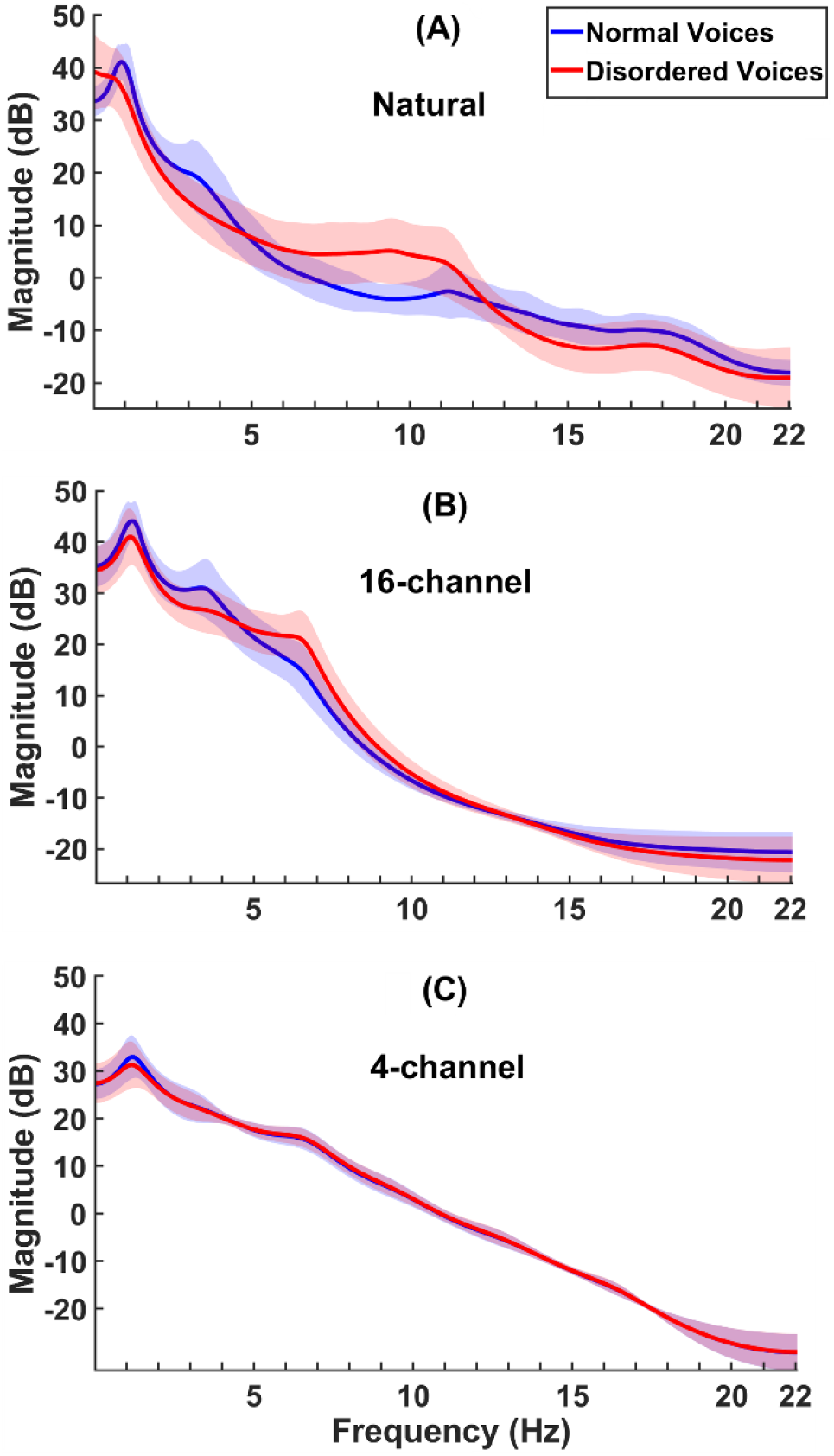
(Color online) Average magnitude spectra of voice stimuli with normal (blue or dark gray lines) and disordered (red or light gray lines) qualities for (A) unprocessed voices, (B) simulated cochlear-implant voices with 16-channel, and (C) 4-channel in the CI noise-vocoder. The standard deviations of the magnitude spectra are also shown as the blue (dark gray) shaded area across the average blue line (dark gray) for the group of normal voices and as the red (light gray) shaded area across the average red line (light gray) for the group of disordered voices.

The difference between the average magnitude spectrum for normal voices (the blue or dark gray line) and that of disordered voices (the red or light gray line) in Fig. 1A reveals distinctive patterns of spectral energy within low-, mid-, and high-frequency regions. For the unprocessed condition, a peak in the frequency regions between 1 and 2 kHz distinguishes average spectrum of voices with normal quality from the average spectrum of disordered voices. These degraded low-frequency patterns in the disordered voice spectrum are considered to be signs of partial closure of the vocal folds (Hartl et al., 2001; Kitzing & Åkerlund, 1993; de Krom, 1995). These differences in the spectral level in frequency bands covering the first formant (*e.g*., breakdown in formant structure) may be associated with breathy voice quality reported in some voice disorders (Kitzing & Åkerlund, 1993; Krom, 1995; Rontal, Rontal, & Rolnick, 1975; Thomas, 2008; Wolfe & Bacon, 1971). The relative reduction in low-numbered harmonic components is also visible in low-frequency regions, which is due probably to irregular vibratory patterns of vocal folds and hoarse voices in disordered voice quality (Fex, Fex, Shiromoto, & Hirano, 1994; Roy & Leeper, 1993; Thomas, 2008; Yanagihara, 1967). A relatively higher level of energy in mid-frequency bands (~4.7 KHz-12.4 kHz) is evident in the average spectrum of samples with disordered voice quality compared to those from the normal group, which potentially signals the presence of high degree of breathiness in disordered voice samples and an increase in the level of the turbulence noise components in the vocal excitation signal (Askenfelt & Hammarberg, 1986; Fukazawa et al., 1988; Hanson, 1997; O’Leidhin & Murphy, 2005). The presence of a wide-band noise in this frequency region (i.e., between ~ 5 kHz and ~12 kHz) in the average spectrum of disordered voices may also be attributed to a rough voice quality in disordered voices (de Krom, 1995).

Comparing the average spectrum of two groups of voices with normal and disordered qualities among three levels of spectral degradation in Fig. 2 (unprocessed in panel (A), 16-channel in panel (B), and 4-channel in panel (C)) demonstrates that CI noise vocoding substantially degrades acoustic information involved in voice quality distinction. In general, the noise vocoding process-simulating CI speech processing - caused a major loss of acoustic information that might be used by listeners to distinguish voice quality at low-, mid-, and high-frequency ranges of voice spectra. The detrimental effect of noise-vocoding increased as the number of channels decreased; this pattern was observed to the extent that spectra of two classes of voice qualities become almost visually indistinguishable at 4-channel CI-simulated voices. This reduction in acoustic distance between groups of voices with normal and disordered qualities highlights the detrimental effect of low spectral resolution in CI speech processing on discarding voice quality-related acoustic information. As the spectra of normal and disordered voices for 16- and 4-channel noise-vocoded voices suggest, a large portion of spectral information at low frequencies is expected to be discarded due to CI processing. This spectral region is particularly important for the perception of voice quality variations as it is where the low-numbered harmonics are located. CI listeners do not have access to the frequency components in these low-frequency regions (Bernstein & Oxenham, 2003; Smurzynski, 1990), which may negatively impact their ability to perceive variations in voice quality. As the number of spectral channels decreases, the noise-like distinctive patterns in the mid-frequency range (between 5 and 12 kHz) disappear, suggesting that listeners with CIs do not have access to acoustic cues relevant to variations in voice quality within these spectral regions.

We further quantified the acoustic distance between voices with normal and disordered qualities at seven levels of spectral degradation to examine the effects of simulated CI processing on acoustic information distinctive of voice quality. Fig. 3 illustrates an example of the filterbank of mel-spaced triangular filters through which voice spectrum of each normal or disordered voice was passed to characterize the variations in their spectral energy at different frequency sub-bands.

**Fig. 3.**
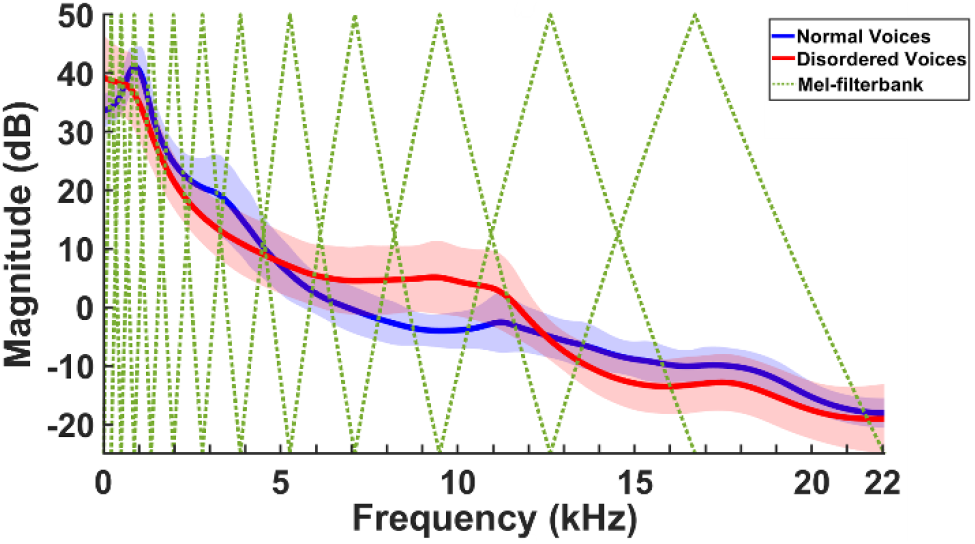
(Color online) The filterbank of mel-spaced triangular filters (green or dark gray dotted lines) superimposed on average magnitude spectra of voices with normal (blue or dark gray line) and disordered (red or light gray line) qualities. In this example, mel-filterbak contains 12 filters, which starts at 0 Hz and expands to 22.05 kHz, corresponding to half of the sampling frequency (44.1 kHz). The actual filterbank in the calculation of MFCC features was comprised of 32 mel filters.

Fig. 4 shows the calculated MD between MFCCs of normal and disordered voices as a function of different levels of spectral degradation, corresponding to change in the number of spectral channels in CI-simulated voices. This figure illustrates that the acoustic distance between voices with normal and disordered qualities decreased largely as a function of noise vocoding, supporting the contention that CI-related processing is detrimental to the reception of voice quality-related information. On average, there was an approximately 33% decline in MD due to noise-vocoding process when comparing the MD at the baseline (i.e., unprocessed condition) (the top dashed line in Fig. 4) with the average MD derived across six levels of spectral degradation (the middle dotted line in Fig. 4). This large decline in acoustic distance between normal and disordered voice qualities suggests that the CI potentially discards an important portion of acoustic information responsible for signaling variation in talkers’ voice qualities.

**Fig. 4.**
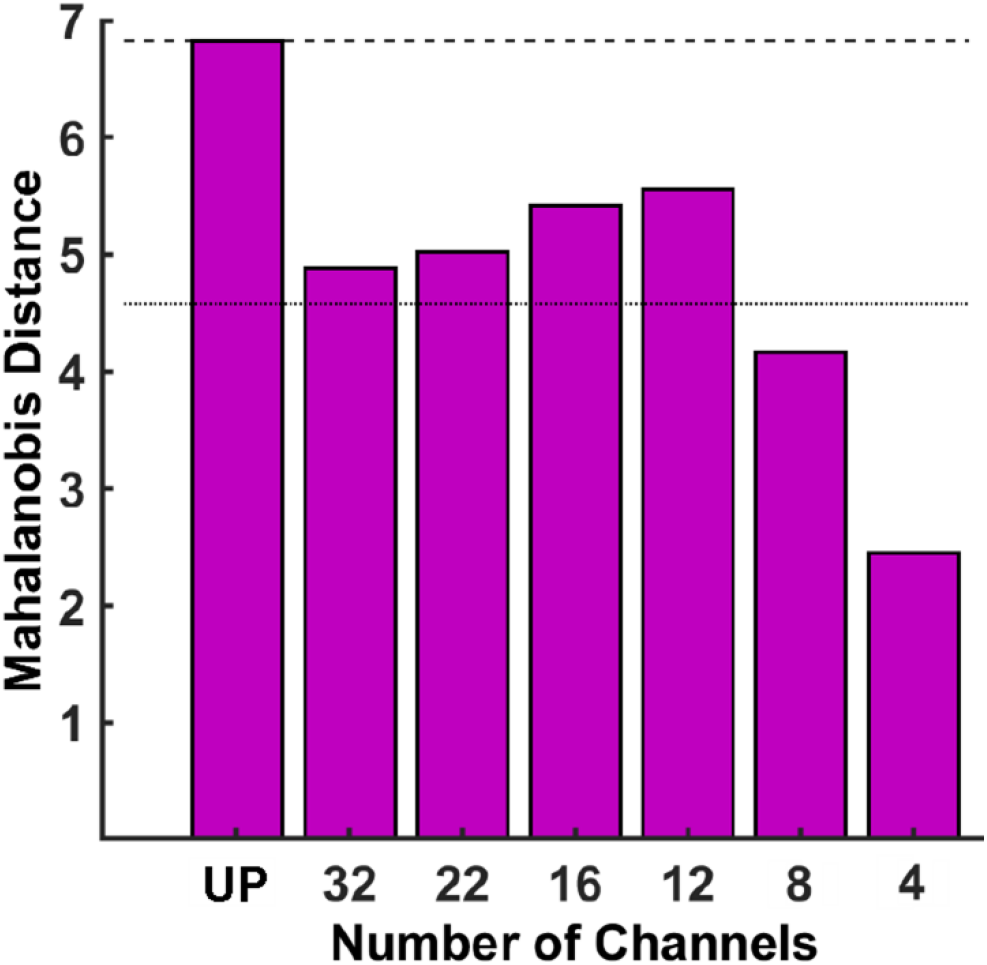
(Color online) *Mahalanobis distance* between two groups of voices with normal and disordered qualities as a function of spectral degradation in CI-simulated voices (i.e., number of spectral channels in noise-vocoder). The top, horizontal dashed line shows the MD derived from unprocessed voices and the dotted line in the middle shows the average of MDs across six levels of spectral degradation. The unprocessed condition is labeled as “UP”.

An interesting pattern was the increase in MD as the number of spectral channels decreased from 32 channels to 22, 16, and 12 channels. Our visual investigation of normal and disordered voice spectra showed that noise vocoding interestingly resulted in more distinctive patterns of spectral energy between normal and disordered voice qualities as the number of spectral channels changed from 32 to 12 channels. This pattern was particularly noticeable in low frequency regions where mel-filterbank is more sensitive to variations in voice spectrum because of its narrower filters, as compared with high-frequency regions with wider filters (see Fig. 3). This unexpected pattern suggests that increasing spectral channels in noise-vocoder does not necessarily mitigate the information loss relevant to voice quality distinction. The range of decrease in acoustic distance (i.e., MD) due to noise vocoding relative to acoustic distance in the unprocessed condition was between ~64% for 4-channel CI-simulated voices and ~19% for 12-channel CI-simulated voice, which was still relatively high.

Fig. 5 shows the results of normal-vs-disordered SVM classification accuracy for seven levels of spectral resolution from the unprocessed condition to the highly spectrally-degraded CI-simulated voices created by 4-channel noise-vocoder. Results from SVM classification supports the general trend displayed by MD on the effect of CI speech processing on acoustic information involved in normal-vs-disordered voice distinction. However, there was an approximately 8% decline in the accuracy of SVM in classification of normal and disordered voice qualities between the unprocessed condition and the average accuracy obtained across six levels of spectral degradation. Classification accuracy in Fig. 5 shows three categories of performance between 80-85%, 85-90%, and 90-95%. These simulated results suggest that the performance of the current CI technology falls within the second category in terms of the effect of number of spectral channels on voice quality distinction (12 channels in MED-EL devices, 16 channels in Advanced Bionics devices, and 22 channels in Cochlear). It is noticeable that the performance in this region varies between 87% and 90% classification accuracy for 12, 16, and 22 channels, which is still at least 5% below the unprocessed condition. We speculate that this difference between SVM and Mahalanobis distance in measuring voice quality-related acoustic distinction is because of the exposure phenomenon simulated by SVM as being trained on a subset of data in a supervised fashion. Another explanation could be related to the calculation of MD, which assumes that features have multivariate normal distribution, which might not be necessarily valid for the MFCC features in this study.

**Fig. 5.**
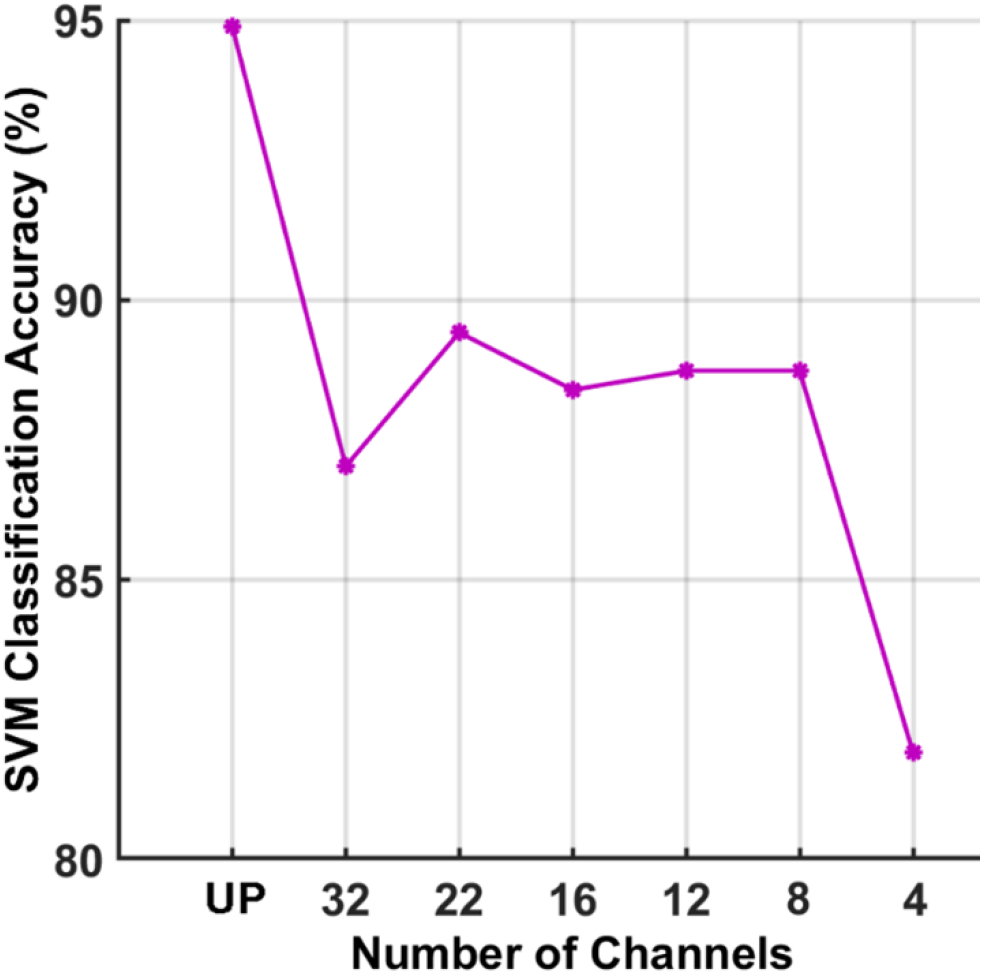
(Color online) The accuracy of SVM in classification between two groups of normal and disordered voices at seven levels of spectral degradation, corresponding to change in the number of noise-vocoder frequency channels (i.e., unprocessed (UP), 32-, 22-, 16-, 12-, 8-, and 4-channels noise-vocoder).

## 4. DISCUSSION AND CONCLUSIONS

This study investigated how CI processing affects acoustic information involved in signaling variations in talkers’ voice quality. We analyzed /a/ vowels spoken by talkers with normal or disordered voice qualities to examine to effects of CI speech processing on acoustic information available for distinguishing voice quality. CI speech processing was simulated by noise-excited envelope vocoders. The results showed large decreases in acoustic distance between normal vs. disordered voices, shedding light on possible difficulties that listeners with CIs may experience in perception of talkers’ voice quality, which in turn may reduce their ability to identify talkers’ voice.

Our investigation of /a/ vowel spectra within different frequency sub-bands across normal vs. disordered voice quality showed that simulated CI processing, based on a noise-vocoder, has a detrimental impact on acoustic information signaling changes in talkers’ voice quality. The CI processing considerably degraded spectral information in low-(<2 kHz), mid-(~4-12 kHz), and high-frequency regions (>12 kHz) of the voice spectrum that could contribute to voice quality distinction (Arjmandi & Pooyan, 2012; Ball & Code, 2008; Behroozmand & Almasganj, 2007). The discriminative spectral information under these frequency regions signals various degrees of distinctive acoustic information that listeners may utilize to perceive fine variations in talkers’ voice quality (Eskenazi et al., 1990; Hillenbrand, Cleveland, & Erickson, 1994; Moisik, 2013; Park et al., 2016; Podesva, 2007; Sicoli, 2010). These patterns of loss in voice quality-related information due to CI processing suggest that listeners with CIs potentially do not receive a large portion of acoustic information, due to the partial transmission of fine-grained spectral structures, that could signal changes in talkers’ voice quality.

We further measured the spectral distance between voices with normal and disordered qualities by first characterizing their vowels’ spectral variations using MFCC features and then calculating the distance between MFCC features using MD. The MD between normal and disordered vowel sounds were examined at different levels of spectral degradation (i.e., unprocessed, 32, 22, 16, 12, 8, and 4-channel CI noise-vocoder processing) to identify how simulated CI speech processing affects the acoustic distance between voices with normal and disordered qualities. We further examined this effect as a function of the number of spectral channels in the noise-vocoder. The results showed a large decrease in acoustic distance between normal and disordered voice qualities because of CI speech processing, highlighting the loss of acoustic information related to talkers’ voice quality in CI processed speech. Therefore, listeners with CIs potentially face difficulties compared to listeners with NH in incorporating talkers’ voice quality information to construct the corresponding mental representation for identification and recognition of talkers’ voice and processing their speech.

The results of SVM accuracy in classification between normal and disordered voice qualities corroborated the detrimental effect of the simulated CI processing on acoustic information involved in voice quality distinction. However, the noise vocoding and the number of channels in the noise-vocoder resulted in lower degrees of drop based on SVM classification compared to the amount of decline quantified by MD. Our interpretation of this difference between SVM and MD is that the training phase in SVM somehow simulated the effect of exposure/learning in making sense of the degraded voice signals for voice quality distinction. The results of SVM classification, as executed through 5-fold cross-validation procedure, suggested a good performance of higher than 80%, even in the highly degraded condition of 4 spectral channels. The average drop of ~ 7% in normal-vs-disordered voice classification between unprocessed and CI-simulated conditions further highlights the detrimental effect of CI speech processing to acoustic information relevant to the recognition of talkers’ voice.

These results can be interpreted in the context of perception of voice cues involved in talker recognition, as well as speech processing. The observed lack of faithful transmission of acoustic information, that is more or less related to various perceptual attributes of voice quality (e.g., breathiness, harshness, and strain), suggests that CI listeners may not benefit from voice quality-related acoustic variations as much as their peers with NH in processing segmental and suprasegmental information for speech comprehension (Dicanio, 2009; Dilley et al., 1996; Dilley et al., 2016; Garellek & Keating, 2011; Gordon, 2001; Gordon & Ladefoged, 2001; Henton, 1986; Ogden, 2001; Redi & Shattuck-Hufnagel, 2001), as well as in recognition of talkers’ gender (Gussenhoven, 2004; Ohala, 1983; Puts, Hodges, Cárdenas, & Gaulin, 2007), race (Alim, 2004; Irwin, 1977; Moisik, 2013; Thomas & Reaser, 2004) and social and cultural class (Esling, 1978; Rilliard et al., 2009; Sicoli, 2007; Stross, 2013; Stuart-Smith, 1999). Our investigation of normal and disordered voice qualities suggests that CI processing substantially degrades spectral properties signaling voice quality variations (Dicanio, 2009; Garellek & Keating, 2011), which probably negatively impact CI listeners’ access and learning talker-specific information as an important skill for robust speech recognition (Johnson, 2005; Kleinschmidt & Jaeger, 2015; Pisoni, 1992). Results from prior studies demonstrated that listeners with CIs do not have access to low-numbered harmonics for robust perception of *F_0_*, leading to poor performance in talker identification and discrimination (Gaudrain & Baskent, 2018), prosody perception elicited by dynamic pitch (Deroche, Kulkarni, Christensen, Limb, & Chatterjee, 2016), and speech recognition in complex listening conditions such as multi-talker situations (Rosen, Souza, Ekelund, & Majeed, 2013; Stickney, Assmann, Chang, & Zeng, 2007; Stickney, Zeng, Litovsky, & Assmann, 2004). In addition, listeners with NH may incorporate other cues such as vocal-tract length (VTL) and formant frequencies in constructing talkers’ voice quality to distinguish between talkers, cues that are poorly perceived by listeners with CIs (Gaudrain & Baskent, 2018). Our results provide further evidence in explaining the poor performance of listeners with CIs in perception and effective use of talkers’ voice cues (Başkent, Luckmann, Ceha, Gaudrain, & Tamati, 2018; Gaudrain & Baskent, 2018; Mehta, Lu, & Oxenham, 2020; Mehta & Oxenham, 2017; Moore & Carlyon, 2005; Stickney et al., 2007) by showing that an important portion of this acoustic information is discarded by cochlear implant speech processing.

There are multiple limitations in the current CI devices including the number of spectral channels (electrodes), which restricts spectral and temporal resolution in CI devices in representation of speech. Our results highlight the need for improving CI speech processing strategies to assure that acoustic cues related to voice quality are faithfully transferred through CIs. Developing more effective strategies requires researchers to evaluate the mechanisms underlying encoding spectral and temporal cues responsible for representing voice quality measures. Therefore, further studies are required to understand how listeners with CIs perceive acoustic cues related to voice quality variations and how possible loss of information at this level may impact their ability to identify talkers and process their speech. Tamati et al., (2017) found that listeners with CIs perform poorer than their NH peers in speech recognition when there is large talker variability. They also showed that CI users experience difficulties in the recognition of talkers’ voices and accents while their performance were also largely variable compared to listeners with NH. Results from the present study provide further evidence that listeners with CIs may not have access to voice quality cues for robust identification of talkers. This lack of access to the voice quality cues may negatively impact CI listeners’ ability to overcome talker variability for successful speech perception.

Despite the limitations of CIs in robust and reliable transformation of speech, Vongpaisal et al. (2010) showed that children with CIs are able to develop models of talker identity, which may reflect the important role of neural plasticity and more powerful speech processing at higher cortical levels for auditory processing and language development. As simulated by SVM, there might be a large effect of exposure or training that can improve the performance of CI listeners in distinction between various voice qualities. In fact, this phenomenon can be logically expanded to how CIs listeners may use the information in the voice delivered through CI at higher levels of speech processing and language learning to compensate for the lack of various acoustic cues such as those related to the perception of talkers’ voice quality (Moore & Shannon, 2009). Speech recognition of children with CIs significantly improved as they had more experience in listening to speech through a CI device (Brown et al., 2004; Fryauf-Bertschy, Tyler, Kelsay, Gantz, & Woodworth, 1997; Miyamoto, Osberger, & Kessler, 1996; Tyler et al., 2000). Another factor that is not modeled in our study is the effect of linguistic and contextual cues in continuous speech that listeners with CIs can incorporate to infer talkers’ voice quality for talker recognition and language processing. The significant effect of these cues on sentence recognition was shown in listeners with CIs (Geers, 2002; Meyer & Svirsky, 2000). Despite these contextual effects, having access to acoustic information relevant to talkers’ voice quality is still critical for speech processing (Başkent et al., 2018; Gaudrain & Baskent, 2018), particularly in complex listening conditions such as speech recognition in multi-talker scenarios (Rosen et al., 2013; Stickney et al., 2007, 2004).

The present study had some limitations that should be considered while interpreting the findings. Although studies based on CI-simulated speech are valuable in general, these findings should be viewed as a general trend rather than the actual performance of listeners with CIs in perception of talkers’ voice quality. One factor is the place-frequency mismatch due to the configuration of the electrode array in the cochlea that its effect is not examined in this study. Furthermore, characterization of voice quality based on MFCCs features might not completely reflect the normal hearing system in perception of voice quality variations as shown by recent studies (Anand, Kopf, Shrivastav, & Eddins, 2019; Eddins, Anand, Lang, & Shrivastav, 2020). It is also worth mentioning that listeners may incorporate segmental and suprasegmental cues at word and/or sentence levels for recognition of talkers’ voice quality rather than merely relying on spectral variations of vowel sounds. Regardless of these limitations, the present study provided new evidence showing that acoustic information involved in distinguishing talkers’ voice quality is substantially degraded in CI-simulated voices. Our results suggest that listeners who use CIs may have great difficulties incorporating voice quality cues for talkers’ voice recognition. The poor spectral resolution provided by cochlear implant device to CI listeners may negatively impact acoustic cues involved in voice quality transmission, leading possibility to subsequent poor perception of talkers’ voice quality in listeners with CIs. Future perceptual studies will determine which specific acoustic cues relevant to talkers’ voice quality are not faithfully transmitted through cochlear implants. The findings from the current study underscore the need in two directions: (a) the need for examining the current signal processing strategies in CIs for their fidelity in passing voice quality cues and developing more advanced speech processing strategies in CI device to assure faithful transmission of these cues, and (b) the need for active use of multimodal (i.e., gesture, tactile, and visual) communicative behaviors to provide supportive cues for listeners with CIs in recognition of talkers’ voice.

## Data Availability

The MEEI Voice Database is property of Kay Elemetrics Corporation and needs to be ordered through this company.

## Acknowledgments

This work was partially supported by Dissertation Completion Fellowship, awarded by Michigan State University to Meisam K. Arjmandi.

## Notes

### Competing Interest Statement

The authors have declared no competing interest.

### Funding Statement

No external funding was received for this study.

### Author Declarations

The data was from Voice Disorders Database model 4337, developed by Massachusetts Eye and Ear Infirmary (MEEI), Voice and Speech Lab.

